# Suicide after leaving the UK Armed Forces 1996-2018: a cohort study

**DOI:** 10.1101/2022.12.12.22283340

**Authors:** Cathryn Rodway, Saied Ibrahim, Jodie Westhead, Lana Bojanić, Pauline Turnbull, Louis Appleby, Andy Bacon, Harriet Dale, Kate Harrison, Nav Kapur

## Abstract

**Background:** There are comparatively few international studies investigating suicide in military veterans and no recent UK studies. We aimed to investigate the rate, timing, and risk factors for suicide in personnel who left the UK Armed Forces (UKAF) over a 22-year period.

**Methods and findings:** We conducted a retrospective cohort study of suicide in personnel who left the regular UKAF between 1996 and 2018 by linking national databases of discharged personnel and suicide deaths. Of the 458,058 individuals who left the UKAF, 1,086 (0.2%) died by suicide. The overall rate of suicide in veterans was not greater than the general population (SMR [95% CI] 94 [88-99]). However, suicide risk was two to four times higher in male and female veterans aged under 25 years than in the same age groups in the general population (age-specific mortality ratios ranging from 160 to 409). Male veterans aged 35 years and older were at reduced risk of suicide (age-specific mortality ratios 47 to 80). Male sex, Army service, discharge between the ages of 16 and 34 years, being untrained on discharge, and length of service under 10 years were associated with increased suicide risk. Factors associated with reduced risk included being married, a higher rank and deployment on combat operations. The rate of contact with specialist NHS mental health services (273/1,086, 25%) was lowest in the youngest age groups (10% for 16–19-year-olds; 23% for 20–24-year-olds).

**Conclusions:** Suicide risk in veterans is not high but there are important differences according to age, with higher risk in young men and women. We found a number of factors which increased the risk of suicide but deployment was associated with reduced risk. Our focus should be on improving and maintaining access to mental health care and social supports for young service leavers, as well as implementing general suicide prevention measures for all veterans.

## INTRODUCTION

Suicide accounts for around 700,000 deaths per year across the world [1] and its prevention is a global health priority [1]. Although suicide rates are generally lower in people serving in the Armed Forces than in the general population [2-4], data suggest the number of suicide deaths by male army personnel in the United Kingdom (UK) and United States (US) has increased in recent years [3,4]. Despite considerable policy interest in the mental health of veterans [5], research investigating suicide in this group is relatively sparse and often conflicting. In the US, suicide rates for veterans peaked in 2018 but have since fallen [6]. Studies in the US have also shown veterans to have a higher rate of suicide compared to the US general population [6-8]. Similar patterns have been reported in Australian and Canadian veterans, with higher suicide rates reported in female veterans, in particular, compared to the female general population [9,10]. In male veterans of the Canadian Armed Forces suicide risk has been consistently higher than the Canadian general population since 1976, although it has fallen, albeit not significantly, in more recent years [10]. Retrospective cohort studies of UK, Dutch, and deployed Swedish veterans, however, report no overall increase in suicide risk compared with their respective general populations [11-14].

There is a lack of global consensus on whether past military service increases subsequent suicide risk, but individual studies have investigated factors which may be associated with veterans’ suicide. Increased suicide risk has been reported in young (24 years and under) male veterans, older (over 40 years) female veterans, veterans with depression or alcohol problems, those subject to early discharge from the Armed Forces, and those who left service more than 20 years previously [11,12,15-18]. The literature also suggests that veterans of the Armed Forces may be a potentially vulnerable group because of adverse life events prior to enlisting, childhood trauma, the difficulties associated with the transition to civilian life, high rates of unemployment following discharge, homelessness, and alcohol and drug misuse [17-22]. Evidence on the link between deployment and suicide risk is mixed with some US studies reporting increased risk in those who were deployed [23,24] and others reporting reduced risk [7].

There have been few systematic investigations of suicide in UK veterans. We have previously published the findings of a 10-year retrospective study of suicide in a cohort of almost 234,000 individuals who had left the UK Armed Forces between 1996 and 2005 [12]. In this cohort, 224 (0.1%) veterans died by suicide. Although we found the overall rate of suicide was not greater than that in the general population, the risk of suicide in young male veterans was elevated. A more recent study of nearly 80,000 Scottish veterans has also found no overall increase in suicide risk compared to the wider population, although the study did report an increased suicide risk in older female veterans with suicide most common in mid-life [11].

Since the publication of our previous study [12], the military context in the UK has changed. A period of intensive operational activity has ended (e.g. Iraq, Afghanistan), with related concern about the mental health impact of having served in these conflicts [25]. The number of full-time personnel in the Army has decreased (by 19% since 2012) [26] and the age distribution has also changed with fewer recruits aged under 18 years [27]. A range of new community-based NHS mental health services for veterans, collectively named “Op COURAGE”, in conjunction with the creation of a specific body within the NHS with Armed Forces expertise [28], may have improved access to mental health care in England. The devolved administrations have their own mental health support provision for veterans, e.g., the Veterans First Point Service in Scotland. Third sector organisations also continue to raise awareness and deliver support and services to those who have left the UK Armed Forces [29,30]. The Government’s Armed Forces Covenant, published in 2011 [31], has also pledged to treat those who serve or have served in the UK Armed Forces fairly and introduced the requirement within the NHS Constitution [32] that “veterans are not disadvantaged in accessing health services”. There is potentially greater public awareness and reduced stigma associated with mental health problems in general [33]. In the general population, there has been a shift in the pattern of age-related suicide rates; men aged 45-64 years are the group at highest risk of suicide, where previously (before 2010) it was younger men aged 25-44 years [34].

Our previous study of suicide in UK veterans provided data to the end of 2005 [12]. In the current study we include over 200,000 additional veterans and an additional 13 years of data. This has enabled a more comprehensive and contemporaneous examination of the burden of suicide in UK veterans and allowed us to identify key characteristics that could inform preventive efforts. The specific aims of this study were to: (i) investigate age-specific rates of suicide in veterans and compare these with rates in the serving and general populations; (ii) identify risk factors, characteristics, and service contacts among veterans; and (iii) describe trends in suicide rates in veterans and compare these with trends in the serving and general populations.

## METHOD

### Study design

In this retrospective cohort study of personnel who left the UK Armed Forces (UKAF) our main outcome was death by suicide after leaving service. We linked national databases of discharged UKAF personnel and deaths by suicide. Comparisons were made with both the general population and serving personnel. We conducted a case control analysis on a subset of this cohort identified as being in contact with mental health services in the 12 months prior to death.

### Study setting and individuals

This study covered England, Scotland, and Wales (due to lack of availability of suicide data from Northern Ireland). Individuals who had had left any of the three branches of the UKAF (Royal Navy (including Royal Marines), Army, and Royal Air Force (RAF)) between 1 January 1996 and 31 December 2018 were included. There was no restriction on length of military service; we included anyone who left after their first day of basic training. The focus of this study was regular UKAF personnel. Individuals who had only served as reserves were excluded from the study because their experiences of service would have differed from those of regular personnel and their characteristics may reflect the general population more closely than they did the serving population [35]. Suicide risk in reservists will be examined in future work. “Other” assignment types such as cadet forces and military provost guard services (MPGS) were also excluded. Regulars included any individual who had been a regular of the UKAF at any point in their service record regardless of their assignment status at last discharge.

### Armed Forces Databases

#### Service Leavers Database

The MOD has a database of all service personnel who have left the UKAF since the mid-1970s (circa 2.1 million records) compiled from the MoD’s Joint Personnel Administration system and legacy pay systems (pre-2005); known as the Service Leavers Database (SLD). Defence Statistics, the team within the MoD responsible for compiling staffing, surveys, and health statistics, compiled an extract from the SLD on all personnel alive on the day of exit for the period 1 January 1996 to 31 December 2018. The starting year was chosen because of the availability of clinical and general population data from that point onwards. The initial extract included a limited number of core variables (name, date of birth, date of exit from Service) which were used for linkage purposes. A further extract of pseudo-anonymised variables was provided once database linkage was complete (see database linkage sub-section below).

### Suicide Databases

We used two databases held by the National Confidential Inquiry into Suicide and Safety in Mental Health (NCISH) to identify deaths by suicide during the study period (1997-2018).

#### General population suicide database

The NCISH general population suicide database included all suicide deaths in the UK. The database is collated from national mortality data on all people who died by suicide obtained from the Office for National Statistics (for deaths registered in England and Wales), National Records of Scotland (for deaths registered in Scotland) and the Northern Ireland Statistics and Research Agency (for deaths registered in Northern Ireland). For this study, data on deaths in Northern Ireland were not included due to restrictions in the disclosure of confidential person identifiable data for health and social care purposes [36]. Individuals who received a conclusion of suicide or intentional self-harm (International Classification of Diseases, Tenth Revision (ICD-10) codes X60-X84) or events of undetermined intent (ICD-10 codes Y10-Y34, excluding Y33.9, Y87.0, and Y87.2), were included in the study sample, as is standard for suicide research [37]. This is because most undetermined deaths are likely to be suicide deaths. These deaths are collectively referred to as suicides in the remainder of this paper. The variables in the NCISH general population database included age, gender, and method of suicide.

#### Database of suicide deaths in people in recent (<=12 month) contact with NHS mental health services

The NCISH patient database is a detailed clinical database of patients who have been in contact with specialist NHS mental health services in the 12 months prior to their death. From national data on all people who died by suicide, mental health providers identified which individuals had contact with NHS mental health services in the 12 months before death. Clinical information was collected via a questionnaire completed by the senior professional responsible for the patient’s care. During the study period, response rates for NCISH questionnaires were 95%. NCISH data collection methods are described in detail elsewhere [38].

### Database linkage

We linked the database of those who had left the UKAF (the SLD) with the NCISH general population suicide database using last name, first name(s)(where available), and date of birth. Of the 1,086 individuals we linked between the two databases, 89% (N=973) were exact matches using all variables, and 4% (N=41) were matched using date of birth, last name, and initials. A further 3,745 individuals matched on date of birth and last name, these were manually checked by members of the research team (SI, CR, JW, LB), equivocal matches were discussed, and 7% (N=72) were verified as veteran suicide deaths based on consensus. Once data linkage was complete, 17 additional pseudo-anonymised variables were obtained from the UKAF databases as described above. These variables included information on demographic characteristics (e.g. age, gender, ethnicity, marital status) and military service (e.g. assignment type (regular/reserve), rank, training indicator, date of entry, length of service, deployment on combat operations (including the Falklands Campaign (1982), Gulf-1 (1990-1991), Iraq (2003-2011), and Afghanistan (2002-2014)), and date and type of discharge (including voluntary and end of contract leavers and involuntary (administrative, disciplinary, and medical) exits). Data were over 95% complete for these variables. The resulting dataset was then linked to the NCISH patient database using a NCISH-derived unique identifier. Names, dates of birth and dates of death, and any unlinked data, were stripped from the database once linkage was complete.

### Statistical analysis

Simple descriptive statistics with numbers and proportions were used to examine the characteristics of former service personnel who died by suicide. Data on ethnicity were excluded due to a high proportion of missing data (82%). We calculated crude suicide rates by time since leaving the UKAF. To compare suicide rates to the UK general population, we calculated age-specific mortality ratios in 5-year age bands and an overall standardised mortality ratio (SMR). To investigate the risk factors that were most associated with suicide by veterans after discharge we used survival analysis, generating hazard ratios (HRs) and 95% confidence intervals (CI) using Cox’s proportional hazards models. The rate of contact with NHS mental health services in the 12 months prior to death was determined using the NCISH patient database. To compare the characteristics of veterans who had died by suicide within 12 months of mental health service contact with individuals in service contact in the general population who died by suicide, a matched case-control design was used. We selected up to 5 controls (general population) per case (former service personnel) matched on age, gender, and year of death from the NCISH patient database. These 1,357 controls had no record of serving in the UKAF. Assuming a prevalence of a risk factor of 30% in the control group and α = 0.05, five controls per case gave us 80% power to examine relative risks of 2.5 and above. Conditional logistic regression analysis was used to explore differences between the cases and controls and unadjusted differences are presented using conditional odds ratios (ORs) and 95% CIs. To compare suicide rates in veterans with the rate in serving personnel, we calculated age-specific mortality ratios in five-year age bands and an SMR using published data on suicide in serving personnel between 1997 and 2018 [3] as a comparator. We do not present age-specific rates for women, as published comparator data were unavailable due to small numbers.

### Ethical approval

The study was approved by the Ministry of Defence Research Ethnics Committee (2071/MODREC/21). Exemption under Section 251 of the NHS Act 2006, enabling access to confidential and identifiable information without informed consent in the interest of improving care, was obtained from the Health Research Authority Confidentiality Advisory Group (21/CAG/0050) and the Public Benefit and Privacy Panel for Health and Social Care (2021-0290). Data were provided by the Healthcare Quality Improvement Partnership (HQIP) from the Mental Health Clinical Outcome Review Programme (MH-CORP) delivered by the National Confidential Inquiry into Suicide and Safety (NCISH)(HQIP362).

## RESULTS

### Characteristics of discharged veterans and those who died by suicide

We obtained data on a total of 458,048 veterans who served as regulars and had left the UKAF between 1^st^ January 1996 and 31^st^ December 2018. These individuals accumulated a total of 5,852,124 person years at risk and a median length of follow-up (interquartile range (IQR)) of 12.9 years (7-18 years). The cohort was predominantly male (416,254, 91%) and the median age (IQR) of the cohort at last discharge from the UKAF was 26 years (21-37 years). Overall, 283,111 (62%) had served in the Army, 88,664 (19%) in the Naval Service, and 86,273 (19%) in the Royal Air Force (RAF). Overall, 38,722 (8%) had a medical reason for discharge.

Based on linkage between NCISH and MoD databases, 1,086 (0.2%) individuals who had served as regulars were found to have died by suicide after leaving the UKAF. Their median age (IQR) at discharge was 23 years (19-31 years); their median age (IQR) at death was 32 years (26-42 years). The majority (1,046, 96%) were male. Around 19% (n=203) of the 1,086 veterans who died by suicide were aged under 25 years, 63% (n=682) were aged between 25-44 years, and 19% (n=201) were 45 years or older. Overall, 799 (74%) had served in the Army, 165 (15%) in the Naval Service, and 122 (11%) in the RAF. The most common method of suicide was hanging or strangulation (n=672, 62%), followed by self-poisoning (n=155, 14%). Firearm deaths were rare (n=27, 2%). Hanging or strangulation was more common in veterans than in the general population during this time period (62% versus 44%; p<0.001) and self-poisoning less common (14% versus 24%; p<0.001). Firearm deaths were similar to the general population (2%; p=0.31). 502 (46%) of the 1,086 veterans who died by suicide were early service leavers (with less than 3 years of service). Of these 502 individuals, 165 (33%) were aged 24 years and younger.

### Rate of suicide

Table 1 shows the age-specific mortality ratio and the SMR for men who left the UKAF compared to the general population. The risk of suicide for male veterans was similar to the risk of suicide in the age-matched general population. If anything, the SMR and 95% CIs suggested the veterans’ suicide rate was very slightly (but statistically significantly) lower than the rate in the general population (SMR [95% CI] 94 [88-99]). However, the risk of suicide for the two youngest veteran age groups (16-19 and 20-24 years) was approximately two to three times higher than their counterparts in the general population (age-specific mortality ratio [95% CI] of 160 [136-187] in those aged 20-24 years and 305 [220-422] in those aged 16-19 years). For men aged over 35 years the age-specific mortality ratios suggested the risk of suicide was lower than for age-matched groups in the general population. Overall, the suicide rate appeared to fall with age (with a significantly increased risk of suicide at ages 16-24, followed by an increased, but non-significant, risk at ages 25-35, then a significantly lower risk from ages 35 to 59).

**Table 1:** Numbers, crude rates per 100,000 person years and age-specific rate ratios for suicide in male regulars who left the UKAF, 1996-2018.

| Age at death (years) | Number of suicide deaths | Crude rate | Age-specific rate-ratio<sup>a</sup> (95% CI) | SMR<sup>b</sup> (95% CI) |
| --- | --- | --- | --- | --- |
| **16-19** | 36 | 28.0 | 304.5 (219.7-422.2) | - |
| **20-24** | 156 | 27.5 | 159.6 (136.4-186.7) | - |
| **25-29** | 198 | 22.2 | 111.0 (96.6-127.6) | - |
| **30-34** | 207 | 22.9 | 102.8 (89.7-117.8) | - |
| **35-39** | 129 | 18.4 | 80.1 (67.4-95.2) | - |
| **40-44** | 121 | 18.1 | 75.2 (62.9-89.9) | - |
| **45-49** | 90 | 15.6 | 66.6 (54.2-81.9) | - |
| **50-54** | 65 | 15.8 | 74.9 (58.7-95.5) | - |
| **55-59** | 25 | 8.8 | 47.2 (31.9-69.8) | - |
| **60-64** | 13 | 9.2 | 62.1 (36.0-106.9) | - |
| **65-87** | 6 | 6.1 | 49.5 (22.3-110.3) | - |
| **16-87<sup>c</sup>** | **1,046** | **19.5** | - | **93.5 (88.0- 99.3)** |

<sup>a</sup> Using the general population as the reference population. Age-specific rate-ratios are expressed so that a ratio under 100 indicates reduced risk and a ratio above 100 indicates an increased risk of suicide.
<sup>b</sup> A SMR indicates the overall risk of this cohort compared to the general population.
<sup>c</sup> Age category up to 87 because no suicide deaths in male veterans serving as regulars occurred in older age groups.

We do not show the breakdown of age-specific data in female veterans due to small numbers. The overall SMR for women who left the UKAF indicated that their risk of suicide was not greater than the risk of suicide in the general population (SMR [95% CI] 126 [92-172]). However, similar to male veterans, there was an increased risk of suicide in the two youngest female age groups compared to the same groups in the general population (age-specific mortality ratios [95% CI] 409 [102–1633] in women aged 16-19 years and 310 [161–596] in women aged 20-24 years). Unlike older male veterans, older female veterans were not at reduced risk but numbers were small.

Suicide rates across services indicated a slightly elevated SMR for men who had served in the Army only (SMR [95% CI] 111 [103-119]), with once again the highest risks in the youngest age groups (age-specific mortality ratios [95% CIs] for males aged 16-19 years: 331 [233-471]; 20-24 years 182 [154-215]; 25-29 years 128 [110-149], and 30-34 years 120 [103-139]). The risk of suicide for men who had served in the Naval Service or RAF was not greater than the risk of suicide in the age-matched general population (SMRs [95% CIs] 69 [59-81] and 61 [51-73], respectively).

### Timing of suicide

Figure 1 shows the rate of suicide by time elapsed since discharge from the UKAF. These data are presented for men only because of the small number of women in individual time period categories. There was some year-on-year variation, but the risk of suicide appeared to be persistent. There was possible evidence of an increase in risk between years 8 and 14 after discharge, which may have reflected the underlying age of the veteran cohort.

**Figure 1:**
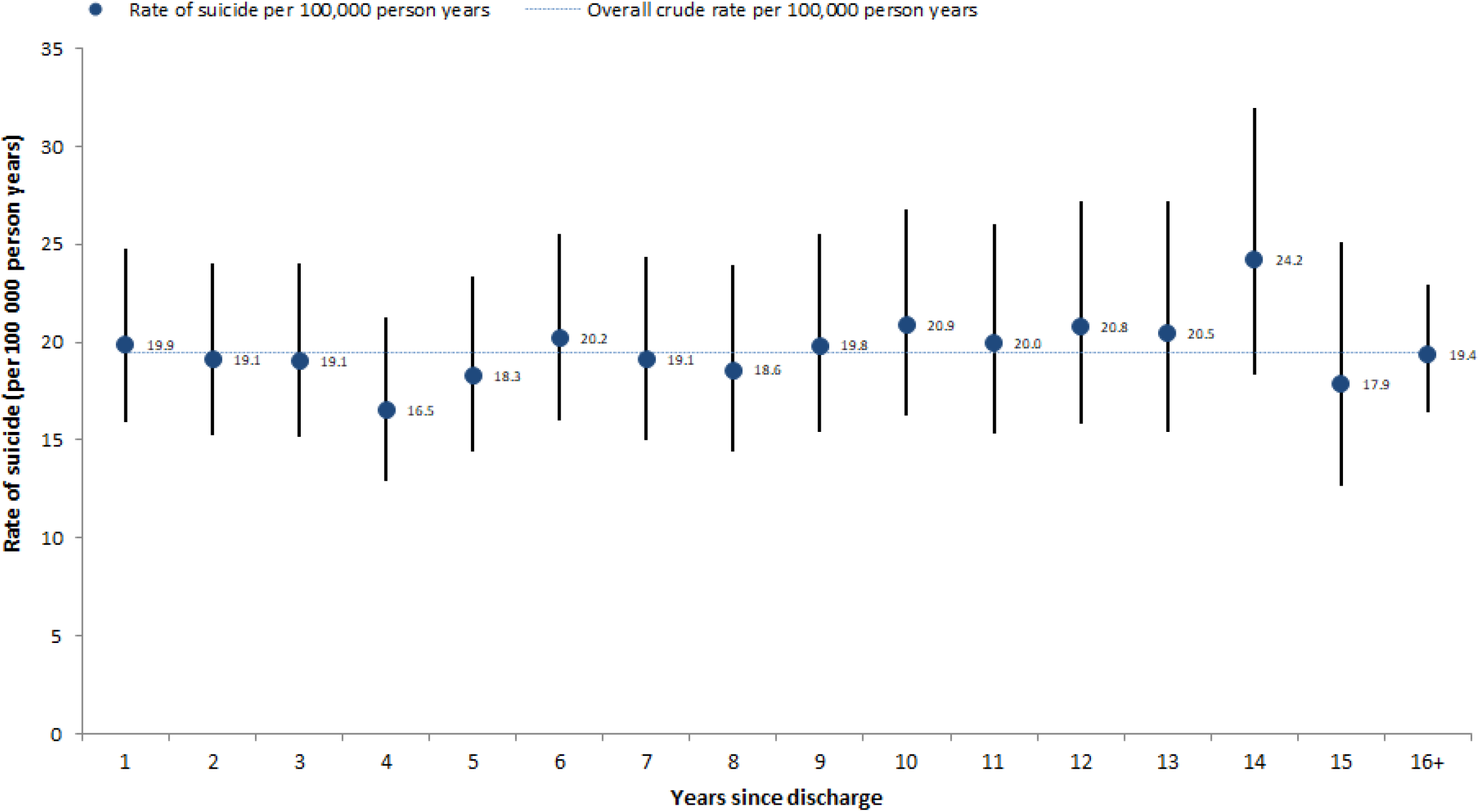
Rate of suicide in male veterans serving as regulars by time elapsed since discharge.

### Risk factors for suicide

An increase in suicide risk was associated with male sex, being discharged from the UKAF between the ages of 16-34 years, being untrained on discharge, having served for less than 10 years, and receiving an administrative, disciplinary, or medical discharge (Table 2). Being married, being an officer (including a non-commissioned officer) and deployment on combat operations were associated with reduced suicide risk (Table 2). Serving in the Navy and the RAF were also associated with reduced suicide risk compared to serving in the Army.

**Table 2:** Risk factors for suicide.

| **Variable** | **Categories** | **N (at risk)<sup>a</sup>** | **Number dying by suicide** | **HR<sup>b</sup> (95% CI)** | ***p*-Value** |
| --- | --- | --- | --- | --- | --- |
| **Age (y) at service exit** | 16-19 | 75,503 | 287 | 2.62 (1.97-3.49) |  |
|  | 20-24 | 114,562 | 321 | 2.25 (1.69-2.98) |  |
|  | 25-29 | 87,914 | 180 | 1.75 (1.30-2.37) |  |
|  | 30-34 | 51,354 | 93 | 1.43 (1.03-1.99) |  |
|  | 35-39 | 38,602 | 70 | 1.20 (0.84-1.71) |  |
|  | 40-44 | 49,662 | 79 | 1.18 (0.84-1.67) |  |
|  | 45+ | 40,451 | 56 | Base | <0.001 |
| **Gender** | Male | 416,254 | 1,046 | Base | <0.001 |
|  | Female | 41,794 | 40 | 0.37 (0.27-0.51) |  |
| **Service** | Army | 283,111 | 799 | Base | <0.001 |
|  | Navy | 88,664 | 165 | 0.61 (0.51-0.72) |  |
|  | RAF | 86,273 | 122 | 0.52 (0.43-0.63) |  |
| **Rank** | Non-officer rank | 252,299 | 761 | Base | <0.001 |
|  | Officer | 44,659 | 41 | 0.28 (0.20-0.38) |  |
|  | NCO | 160,675 | 271 | 0.51 (0.44-0.58) |  |
| **Marital status** | Single | 248,231 | 727 | Base | <0.001 |
|  | Married | 139,918 | 227 | 0.51 (0.44-0.59) |  |
|  | Divorced/separated | 7,681 | 10 | 0.72 (0.39-1.35) |  |
| **Training<sup>c</sup>** | Trained | 270,112 | 579 | Base | <0.001 |
|  | Untrained | 141,363 | 404 | 1.27 (1.12-1.44) |  |
| **Type of discharge** | Normal | 82,215 | 134 | Base | <0.001 |
|  | Disciplinary | 26,252 | 51 | 2.67 (1.93-3.69) |  |
|  | Medical | 38,722 | 122 | 2.55 (1.99-3.26) |  |
|  | Administrative | 86,924 | 357 | 2.88 (2.36-3.52) |  |
|  | Own request | 107,320 | 191 | 1.36 (1.09-1.69) |  |
| **Deployed on combat operation<sup>d</sup>** | No | 328,875 | 899 | Base | <0.001 |
|  | Yes | 129,173 | 187 | 0.71 (0.60-0.83) |  |

**Table 2 (continued): Risk factors for suicide**
| Variable | Categories | N (at risk)<sup>a</sup> | Number dying by suicide | HR<sup>b</sup> (95% CI) | *p*-Value |
| --- | --- | --- | --- | --- | --- |
| Length of service (y) | <1 | 96,653 | 328 | 2.18 (1.82-2.61) |  |
|  | 1 to <2 | 22,790 | 98 | 3.11 (2.43-3.97) |  |
|  | 2 to <3 | 16,230 | 76 | 3.41 (2.61-4.45) |  |
|  | 3 to <4 | 21,798 | 60 | 1.87 (1.40-2.50) |  |
|  | 4 to <5 | 35,705 | 71 | 1.59 (1.21-2.09) |  |
|  | 5 to <10 | 104,455 | 203 | 1.51 (1.23-1.84) |  |
|  | 10 to <15 | 45,245 | 65 | 0.96 (0.72-1.27) |  |
|  | 15+ | 115,172 | 185 | Base | <0.001 |

<sup>a</sup> Numbers for each category may not tally with the total number at risk due to missing data;
<sup>b</sup> Unadjusted HRs calculated using Cox's proportional hazards modelling. Males and females both included. Total number at risk 458,048, total number of suicide deaths 1,086;
<sup>c</sup> Trained refers to individuals who have completed phase 1 (basic) and phase 2 (skill-based) training;
<sup>d</sup> Includes deployment to Falklands Campaign (1982), Gulf-1 (1990-1991), Iraq (2003-2011), and Afghanistan (2002-2014).
NCO = Non-commissioned officer

### Rates and characteristics of veterans in contact with mental health services prior to suicide

Of the 1,086 individuals who died by suicide after leaving the UKAF, 273 (25%, 95% CI 22–28) had been in contact with NHS mental health services in the 12 months prior to their death. This is similar to the proportion of individuals in contact with NHS mental health services in the 12 months before suicide in the general population (27%). The proportion of individuals who had left the UKAF and were mental health patients prior to suicide was lowest in the youngest age groups, although numbers were small (10% in those aged 16-19 years; 23% in those aged 20-24 years).

Table 3 compares the characteristics of veterans who died by suicide within 12 months of contact with NHS mental health services (cases) with individuals matched by age, gender, and year of death, who also died by suicide within 12 months of contact with NHS mental health services but had not served in the UKAF (controls). There were few differences between cases and controls. Veterans of the UKAF who died by suicide were more likely to have a primary diagnosis of affective disorder (depression or bipolar disorder), evidence of a post-traumatic stress disorder (PTSD) diagnosis (including a primary, secondary, tertiary, or quaternary diagnosis, although numbers were small), and to have had their first contact with NHS mental health services within the 12 months prior to death. They were less likely to have been on long-term sick leave at the time of death and to have a diagnosis of schizophrenia and other delusional orders.

**Table 3:** Characteristics of individuals who left the UKAF and who had contact with mental health services in the 12 months prior to suicide and matched controls who had not served in the UKAF.

| Risk group | Variable | Cases (n=273)<sup>a</sup> | Controls (n=1,357)<sup>a</sup> | p-Value<sup>b</sup> | OR (95% CI) |
| --- | --- | --- | --- | --- | --- |
| **Demographic features** | Unmarried | 200 (78%) | 1,053 (81%) | 0.21 | 0.81 (0.58-1.12) |
|  | Unemployed | 156 (61%) | 758 (61%) | 1.00 | 1.00 (0.76-1.32) |
|  | Long-term sick | 19 (7%) | 157 (13%) | **0.02** | **0.57 (0.35-0.95)** |
|  | Living alone | 121 (48%) | 602 (48%) | 0.82 | 1.03 (0.78-1.36) |
|  | Homeless | 10 (4%) | 60 (5%) | 0.72 | 0.88 (0.44-1.77) |
| **Service characteristics** | In-patient | 21 (8%) | 108 (8%) | 0.84 | 0.95 (0.58-1.55) |
|  | Recent (< 3 months) discharge | 38 (15%) | 193 (16%) | 0.91 | 0.98 (0.66-1.44) |
|  | Under CR/HT services | 30 (12%) | 114 (9%) | 0.15 | 1.39 (0.90-2.15) |
|  | Missed last contact | 73 (30%) | 357 (30%) | 0.69 | 1.06 (0.78-1.44) |
|  | Non-adherent with medication | 29 (13%) | 173 (15%) | 0.64 | 0.90 (0.59-1.39) |
| **Clinical characteristics** | Schizophrenia & other delusional disorders | 41 (15%) | 325 (25%) | **0.0006** | **0.55 (0.39-0.79)** |
|  | Affective disorders (depression or bipolar disorder) | 90 (33%) | 363 (27%) | **0.04** | **1.35 (1.01-1.80)** |
|  | Alcohol dependence/misuse | 31 (11%) | 129 (10%) | 0.38 | 1.21 (0.80-1.84) |
|  | Drug dependence/misuse | 24 (9%) | 147 (11%) | 0.27 | 0.77 (0.49-1.23) |
|  | Personality disorder | 34 (13%) | 122 (9%) | 0.10 | 1.43 (0.94-2.16) |
|  | Any PTSD diagnosis<sup>c</sup> | 9 (3%) | 14 (1%) | **0.01** | **3.29 (1.37-7.87)** |
|  | Any secondary diagnosis | 154 (57%) | 784 (59%) | 0.56 | 0.92 (0.70-1.21) |
| **Behavioural characteristics** | History of illness < 12 months | 59 (24%) | 226 (19%) | 0.09 | 1.33 (0.96-1.85) |
|  | History of self-harm | 173 (68%) | 831 (64%) | 0.29 | 1.17 (0.87-1.57) |
|  | History of violence | 77 (32%) | 392 (32%) | 0.95 | 0.99 (0.73-1.35) |
|  | History of alcohol misuse | 160 (62%) | 735 (57%) | 0.13 | 1.24 (0.94-1.63) |
|  | History of drug misuse | 133 (52%) | 745 (57%) | 0.14 | 0.80 (0.60-1.08) |
| **Contact with services** | First contact with MHS < 12 months | 89 (36%) | 340 (28%) | **0.015** | **1.45 (1.08-1.95)** |
|  | Last contact < 7 days | 101 (38%) | 569 (43%) | 0.13 | 0.81 (0.62-1.07) |
|  | Long-term risk: low or none | 134 (60%) | 656 (58%) | 0.67 | 1.07 (0.79-1.43) |
|  | Short-term risk: low or none | 190 (81%) | 1,016 (84%) | 0.25 | 0.80 (0.56-1.16) |

<sup>a</sup> Percentages in brackets expressed as a proportion of valid cases. The number of valid cases may vary between variables (risk groups) due to missing data;
<sup>b</sup> Based on $\chi^2$ test; <sup>c</sup> Any PTSD diagnosis includes a primary, secondary, tertiary, or quaternary diagnosis of PTSD.
CR/HT = Crisis Resolution/Home Treatment; PTSD = Post-traumatic stress disorder; MHS = Mental Health Services.

### Comparison with the in-service sample

Table 4 presents the rates of suicide for men who left the UKAF and men who were serving in the UKAF during the period 1997-2018. The age-specific mortality ratios and SMR compare rates of suicide in men who left the UKAF to rates of suicide in male serving personnel. The results suggested the overall risk of suicide was over twice as high in men discharged from the UKAF than those still serving (SMR [95% CI] 257 [241-273]), with elevated risks across all age groups. Published age-specific suicide rates for serving women were not available to calculate an SMR.

**Table 4:** Number and crude rates per 100,000 person years and age-specific rate ratios for suicide in male regulars who left the UKAF and male regulars serving in the UKAF at the time of death, 1997-2018*.

| Age at death (y) | Discharged sample |  | In-service sample | Age-specific rate ratio<br>(95% CI)<sup>b</sup> | SMR (95% CI)<sup>c</sup> |
| --- | --- | --- | --- | --- | --- |
|  | Number of suicide deaths | Crude rate | Average age-standardised suicide rate<sup>a</sup> |  |  |
| **16-19** | 36 | 29.5 | 12.9 | 228.5 (164.8-316.8) | - |
| **20-24** | 155 | 28.8 | 11.4 | 252.7 (215.9-295.8) | - |
| **25-29** | 198 | 23.7 | 9.5 | 249.4 (216.9-286.6) | - |
| **30-34** | 207 | 25.0 | 7.6 | 328.3 (286.5-376.3) | - |
| **35-39** | 129 | 21.0 | 7.7 | 272.1 (229.0-323.3) | - |
| **40-44** | 121 | 20.8 | 10.9 | 191.0 (159.8-228.2) | - |
| **45-54** | 155 | 17.9 | 6.9 | 259.8 (222.0-304.1) | - |
| **16-54<sup>d</sup>** | **1,001** | **22.8** | **9.0** | - | **256.7 (241.3-273.1)** |

\*Numbers differ to Table 1 because of slightly shorter time period and different age categories;
<sup>a</sup>Source: Ministry of Defence, UK Armed Forces suicides: 2021 [3];
<sup>b</sup> Using the in-service population as the reference population. Age-specific rate-ratios are expressed so that a ratio under 100 indicates reduced risk and a ratio above 100 indicates an increased risk of suicide, and compare suicide rates in the discharged (veteran) sample with rates in the serving sample;
<sup>c</sup> A SMR indicates the overall risk of this cohort compared to the serving population;
<sup>d</sup> Age category restricted to 54 years because no in-service suicide deaths in serving male regulars occurred in older age groups.

## DISCUSSION

### Main findings

We found that overall, veterans were not at an increased risk of suicide when compared to the general population. However, the suicide risk was two to four times higher in men and women aged under 25 years who had left the UKAF. The risk was lower in veterans aged 35 years and older compared to the general population. We found that the risk within the first few years of discharge was fairly persistent but may have increased between years 8 and 14 after leaving service. Men, those who had served in the Army, and those with a length of service of less than 10 years were at greater risk of suicide. Other factors associated with an increased risk of suicide were being discharged from services between the ages of 16 and 34 years, being untrained on discharge, and leaving service involuntarily (i.e. receiving an administrative, disciplinary, or medical discharge). We found higher rank, being married, and deployment on combat operations were associated with a reduced risk of suicide. Methods of suicide in veterans were comparable to the general population, although hanging and strangulation were more common (62% versus 44%) and self-poisoning less common (14% versus 24%) in veterans – potentially reflecting the gender composition of the veteran cohort compared with the general population.

A quarter (25%) of veterans who died by suicide had been in contact with specialist NHS mental health services in the 12 months prior to death (i.e., were patient suicides). The rate of contact with these services was lowest in the age-group at the greatest risk of suicide (21% for those aged 24 years or younger). A primary diagnosis of affective disorder was more common and a diagnosis of schizophrenia less common in veterans who were mental health patients (i.e., cases) compared to mental health patients who had not served in the UKAF (i.e., controls). Evidence of a PTSD diagnosis was uncommon but more likely in cases than controls (3% vs 1%). There were few other differences between our cases and controls. Our examination of suicide rates in veterans compared with serving personnel indicated that suicide risk was over two times higher in veterans, with elevated risks across all age groups. The findings from this current study are broadly consistent with our previous study [12] and with others examining suicide in UK [11] and US veterans [18].

### Strengths and limitations

This study is one of the few to investigate suicide in military veterans and includes a large cohort of individuals to have left the UKAF, with up to 23 years follow-up and linkage to multiple databases. Our findings need to be considered in the context of the following limitations. Firstly, information on those who had left the UKAF was obtained from administrative databases and we were limited by the information contained in those and were unable to explore the role of pre-service vulnerabilities, or factors that may have influenced later suicide risk (for example, unemployment, homelessness). These factors will be examined in the next phase of this study where we will undertake an in-depth examination of a sample of veteran suicide deaths using information from coroner records. Secondly, information on contact with support services was only available for those veterans identified as having been in contact with specialist NHS mental health services. This study could not tell us the number of veterans who had been in contact with their GP prior to suicide, attended the Emergency Department, or who had been in contact with other health, social care, or voluntary services including military Departments of Community Mental Health (DCMH). We aim to examine contact with a range of support services in the next phase of this study. Thirdly, we examined suicide and undetermined deaths only. We did not examine other categories of death which may sometimes include possible suicide deaths (e.g., accidental deaths) or investigate wider causes (e.g., drugs and alcohol, natural causes). Such deaths may be important contributors to early mortality, although evidence suggests both Falkland and Gulf veterans have similar rates of death for external causes of injury and lower rates of death from disease than the general UK population, attributed to the “healthy worker effect” [39]. Fourthly, our analysis of clinical characteristics was based on a help-seeking sample of veterans in recent contact with specialist NHS mental health services (a quarter of the total) who may not have been typical of veterans as a whole and may have differed from veterans seeking help from other sources. Fifthly, veterans who had served as reserves were excluded from our analysis. Reservists may not be exposed to the same experiences as regular personnel, although there is some evidence that reservists may have poorer outcomes than regulars following the transition to civilian life [40]. We plan to examine suicide risk in those who served as reserves in a subsequent publication.

### Interpretation of findings

Some previous studies, but by no means all, have suggested a history of deployment is associated with increased suicide risk [23,24]. For some people their experience of military service is a positive one but for others it may be negative. Public perception may be one of individuals traumatised by deployment on combat operations, struggling to settle back into society post-discharge, and not receiving the mental health care they need. While this might be the case for individual veterans, our results could be interpreted as presenting a somewhat different picture overall. Veterans who were younger and had lengths of service of less than 10 years were at greatest risk of suicide after discharge. Being older and longer lengths of service were protective. A significant proportion of veterans had deployed on combat operations but there was no evidence that such deployment (and perhaps, by extension, combat-related exposure) was associated with suicide. In fact, deployment appeared to reduce risk. Other protective factors included being married and having received training, perhaps highlighting the importance of social support and integration. Individuals who had served in the Army and who had left service involuntarily were at higher risk of suicide. A minority of veterans, particularly younger veterans, were in contact with NHS mental health services, although this is true of men generally. A known diagnosis of PTSD was unusual, even in those under mental health care, but of course this was based on clinical assessments and people who did not present to services or patients where relevant symptoms were not recognised would not have been included. Interestingly, among patients who had sought mental health care, veterans had similar characteristics to non-veterans. This study was unable to examine in detail the effect of pre- or post-service vulnerabilities or in-service exposures on suicide risk, or to estimate the relative contribution of each. Unemployment, financial and relationship problems, and pre-service adversity have been reported as common in veterans [19,22], but this is also true in the wider population, particularly among middle-aged men [41]. We plan to examine all potentially important antecedents in the next phase of this study using additional data sources such as coroners’ records.

### Implications

Almost 1 in 25 people aged 16 years and over in England and Wales is a veteran of the UKAF [42]. The highest risk of suicide continues to be in young people who have left the UKAF and, although all veterans regardless of age should be the target for prevention, younger individuals with short lengths of service may have the most pressing needs. Those who have served in the Army and who leave service on a non-voluntary basis are also at higher risk.

Since the publication of our earlier study [12] over a decade ago, there have been considerable improvements to the care available for serving personnel and veterans, including the introduction of specialist veteran-specific mental health services across the UK (e.g. the Transition, Intervention and Liaison Service (TILS), Complex Treatment Service (CTS) and High-Intensity Service (HIS)). The stigma surrounding mental health has also potentially reduced. The availability of care and awareness of veterans’ mental health needs within healthcare services, however, varies, and there remains a lack of knowledge regarding the Armed Forces Covenant within the healthcare system [5]. What we do know is that while support services for veterans may have increased in the UK in the last decade, the efficacy of that support requires further evaluation. The 2021 Census in England and Wales was also the first to ask if people had ever served in the UKAF [42]. As further data from this becomes available a better understanding of the needs of veterans for service providers will develop.

The low rate of contact with specialist NHS mental health services in our study suggests veterans, particularly younger veterans, may not be seeking help. Although this mirrors findings for men in the general population, specific factors in veterans might be related to stigma or a perception that civilians may not understand the issues they are facing or have faced [43]. Evidence suggests it takes an average of four years for veterans to seek mental health support [5,44]. Interventions that encourage help-seeking or campaigns to reduce stigma lack evaluation but may increase engagement in this population. Equipping health services with the knowledge and training to better support the health of veterans and understand their culture and needs, such as through the Veteran Friendly GP Practice Accreditation Programme [45], the Veteran Aware Accreditation for all NHS Trust providers in England overseen by the Veterans Covenant Healthcare Alliance (VCHA) [46], and the Improving Access to Psychological Therapies (IAPT) positive practice guide for working with veterans [47], may continue to improve health outcomes and reduce suicide risk in this vulnerable population.

A quarter of veterans who died by suicide were in contact with NHS mental health services in the 12 months prior to death, this is roughly equivalent to the proportion of suicide deaths by patients in the UK overall [48]. The characteristics of veteran and non-veteran patient suicides were comparable, suggesting the prevention challenge is similar in veterans and the general population, so tackling previous self-harm and alcohol or drug misuse, increasing awareness of the vulnerability of patients who live alone and enhancing social support could be helpful strategies in veterans as they are in the general population. The persistence of suicide risk suggests that prevention may need to take a long-term perspective.

## Supporting information

Supplemental_STROBE checklist

## Data Availability

Data from this study cannot be shared because of information governance restrictions in place to protect confidentiality. Access to NCISH data can be requested via application to the Healthcare Quality Improvement Partnership (www.hqip.org.uk/national-programmes/accessing-ncapop-data/)

## Acknowledgements

We would like to thank Defence Statistics Health, the Armed Forces Team within NHS England, and staff at the National Confidential Inquiry into Suicide and Safety in Mental Health (NCISH) for their help and advice on the study. We thank Dan Stears, Fiona Naylor, and Liz Monaghan (members of Mutual Support for Mental Health Research (MS4MH-R), the patient and public involvement and engagement group at the Centre for Mental Health and Safety, University of Manchester) and Tom Fox, Jo Brettell, and Wayne Palmer for their advisory roles in the study design. We would also like to thank the Healthcare Quality Improvement Partnership (HQIP) for the provision of data from the Mental Health Clinical Outcome Review Programme (MH-CORP) as delivered by the National Confidential Inquiry into Suicide and Safety in Mental Health.

